# Quantification of non-persistent pesticides in small volumes of human breast milk with ultra-high performance liquid chromatography coupled to tandem mass-spectrometry

**DOI:** 10.1101/2020.09.18.20196162

**Authors:** Theresa L. Pedersen, Jennifer T. Smilowitz, Carl K. Winter, Shiva Emami, Rebecca J. Schmidt, Deborah H. Bennett, Irva Hertz-Picciotto, Ameer Y. Taha

## Abstract

Existing methods for the analysis of pesticides in breast milk involves multiple extraction steps requiring large sample and solvent volumes, which can be a major obstacle in large epidemiologic studies. Here, we developed a simple, low-volume method for extracting organophosphates, pyrethroids, carbamates, atrazine and imidacloprid from 100-200 µL of human breast milk. We tested microwave-assisted acid/base digestion and double solvent extraction with 2 or 20 mL of 2:1 (v/v) dichloromethane/hexane, with or without subsequent solid phase extraction (SPE) clean-up. Samples were analyzed by liquid chromatography tandem mass-spectrometry. Analyte recoveries and reproducibility were highest when 100-200 µL milk were extracted with 2 mL of dichloromethane/hexane without subsequent SPE steps. Analysis of 79 breast milk samples using this method revealed the presence of carbamates, organophsphates, pyrethroids and imidacloprid at detection frequencies of 79-96%, 53-90%, 1-7% and 61%, respectively. This study provides a simple, low-volume method for measuring pesticides in human breast milk.

## Introduction

The banning of persistent halogenated pesticides (e.g. dichlorodiphenyltrichloroethane, aldrin/dieldrin, lindane, toxaphene) in the 1970s and 1980s led to massively increased use of non-persistent organophosphate, pyrethroid and carbamate pesticides in agricultural farms, and in and around homes.^1-2^ Although originally considered safer, long-term exposure to non-persistent pesticides has been associated with multiple health problems including neurological defects, cancer and infertility.^3-6^ Exposure during pregnancy has been linked to poor intellectual development, increased risk of atypical neurodevelopment including cognitive impairments that persist throughout childhood, and autism spectrum disorders.^7-11^

Multiple studies have assessed exposure to non-persistent pesticides by measuring their concentrations in blood, or quantifying their metabolites in urine. ^2, 12-15^ Breast milk, however, remains an understudied exposure matrix, despite studies showing the accumulation of organophosphates, pyrethroids and carbamates there.^16-20^ Studying breast milk is important for probing maternal exposure to non-persistent pesticides, and understanding the potential impact of early life postnatal chemical exposures on neurocognitive and behavioral development.

A major analytical challenge in measuring non-persistent pesticides in breast milk is that the methods used are cumbersome and difficult to routinely perform (e.g. in large cohort studies), because per sample, they typically involve the use of large quantities of organic solvent (10-190 mL) and biospecimen (1-10 mL milk),^16-28^ as well as multiple extraction steps (∼5-10).^16-29^ In some cases, the use of a high-pressure extraction system is required,^16^ making it difficult for laboratories that lack the equipment to isolate and measure pesticides. Additionally, official methods by the National Institute for Occupational Safety and Health (NIOSH) are limited to one class of compounds (e.g. organophosphates, Method 5600) or have not been validated on breast milk matrix.^30^ A simplified but comprehensive analytical method covering a broad range of pesticides used on agricultural farms and in and around homes would be valuable in probing infant exposures through breast milk during the first few months of life.

To overcome these analytical challenges, in the present study we developed a simple method for measuring 28 pesticides in 100-200 µL of breast milk using only 4 mL organic solvent. Below, we first describe our unsuccessful attempts to simultaneously isolate all compounds using microwave-assisted extraction in acid or base followed by C18 solid phase extraction (SPE) clean-up to reduce matrix effects caused by lipids, as well as a published pyrethroid extraction method involving a high-volume (20 mL) liquid-liquid extraction with hexane:dichloromethane (2:1), followed by alumina and C18 SPE which yielded poor recoveries.^18^ We then describe the success of using low volume (2 mL) liquid-liquid extraction with hexane:dichloromethane, to simultaneously isolate 28 non-persistent pesticides belonging to the organophosphate, pyrethroid and carbamate classes, as well as atrazine (a triazine) and imidacloprid (a neonicotinoid), which continue to be used across the US.^31^ The method was then used to quantify pesticide concentrations in a cohort of 79 lactating mothers.

## MATERIALS AND METHODS

### Materials

Pesticide analyte solutions were purchased from AccuStandard, (New Haven, CT USA) and class-specific isotopically labelled surrogates were purchased from Cambridge Isotope Laboratories, Inc. (Andover, MA USA). The internal standard, 1-phenyl-ureido3-hexanoic acid (PUHA), was synthesized and provided as a gift, courtesy of Dr. Bruce Hammock (University of California, Davis). 1-cyclohexyl ureido dodecanoic acid (CUDA) internal standard was purchased from Cayman Chemical (Ann Arbor, MI). Extraction solvents were Optima grade and liquid chromatography mobile phase solvents were liquid chromatography-mass spectrometry (LCMS) grade, purchased from Thermo Fisher Scientific (Waltham, MA USA). Acids, bases, and ammonium formate were purchased from Sigma-Millipore (St. Louis, MO USA).

### Participants and breast milk sample collection

Method development was performed on pooled breast milk samples obtained from 25 mothers enrolled in the Markers of Autism Risk in Babies - Learning Early Signs (MARBLES) study.^32^ MARBLES is a prospective cohort study that enrolled pregnant mothers who had a previous child diagnosed with an autism spectrum disorder,^32^ and are therefore carrying another child who is at high risk of developing autism.^33^ Breast milk samples were collected longitudinally during the first year after delivery. Although the MARBLES protocol includes following the younger sibling to 36 months of age, when a definitive diagnosis is made,^32^ the analysis in the present paper was confined to pooled samples from drop-out mothers, whose children were not successfully followed to a final diagnosis.

Upon developing the method (as described below), we measured pesticides in breast milk of 79 healthy women enrolled in the Foods for Health Institute Lactation Study at UCD. Participants were enrolled at 34-38 weeks of gestation and completed detailed health history questionnaires regarding demographics, anthropometrics, pregnancy history, current and prior health history, dietary habits and restrictions, physical activity level, as well as medication and supplementation intake history. Upon delivery of their infants, mothers reported the mode of delivery (C-section vs. vaginal), infant sex, weight, length, and gestational age at birth, and filled out questionnaires regarding their health and the health of their infants, as well as their diet throughout the study.

Participants received lactation support and training on proper sample collection from the study’s lactation consultant. Participants were instructed to write the time and date of breast milk collection on all sample tubes. Breast milk samples were collected in the morning between days 35 and 42 postpartum from 79 subjects, and on day 249 postpartum from 5 subjects, using a modified published method involving milk collection from one breast using a Medela Harmony Manual Breast pump by the participant 2-4 h after complete milk removal.^34^ Participants fully expressed one breast into a bottle, inverted the bottle 6 times, aliquoted 12 mL into a 15 mL polypropylene tube, and subsequently froze the breast milk sample in their kitchen freezer (−20 °C). All breast milk samples were transported from participants to the lab on dry ice and stored at -80 °C until processing.

At 60 days postpartum, participants in the Lactation Study visited the UCD Ragle Human Nutrition Center to provide a fasting blood sample, and heart rate, blood pressure, weight, and height were measured. Reported participant characteristics (education, ethnicity, parity, birth mode and infant gender) are shown in **Supplemental Table 1**. Maternal and infant anthropometrics, which include maternal age, BMI, blood pressure and heart rate and infant gestational age at birth, birth weight and birth height, are shown in **Supplemental Table 2**.

The subject IDs were blinded to the researchers and the samples were prepared and analyzed in a random order.

The University of California, Davis (UCD) Institutional Review Board (IRB) approved all aspects of the MARBLES and UCD Lactation studies and written informed consent was obtained from all participants prior to collection of data or specimens (IRB # 225645, 216198 and 887479)

### Standard solutions

Three individual master mixtures containing either the pesticide analytes, labelled pesticide class surrogates or the CUDA/ PUHA internal standards were dissolved in methanol. Calibration standards in the range of 0.005 to ∼8000nM were made in methanol from the three master mixtures. A pesticide class-specific deuterated surrogate spike solution was also made in methanol at ∼2000nM concentration. An analyte spike solution of unlabeled pesticide standards listed in **Table 1** was made at ∼1000nM. CUDA/PHAU internal standard reconstitution solution was made in methanol at 200nM. All solutions were capped under nitrogen in sealed amber glass vials, and stored at -20°C.

**Table 1.** AB Sciex 6500 QTrap optimized pesticide parameters for analyte parent ion, product ion, retention time, declustering Potential (DCP) and collision energy (CE).

| Analyte Class/Name | Parent Ion | Product Ion | Retention Time | DCP | CE |
| --- | --- | --- | --- | --- | --- |
| ***Carbamates*** |  |  |  |  |  |
| Formetanate HCl | 222.3 | 165.5 | 2.1 | 20 | 22 |
| Oxamyl | 237.3 | 72.1 | 2.13 | 30 | 28 |
| Methomyl | 163.2 | 88.1 | 2.19 | 25 | 13 |
| <sup>13</sup>C2<sup>15</sup>N-Methomyl (carbamate surrogate) | 166.2 | 90.9 | 2.19 | 20 | 13 |
| Bendiocarb | 224.2 | 109.1 | 2.73 | 15 | 25 |
| Propoxur | 210.2 | 168.1 | 2.74 | 20 | 10 |
| Carbofuran | 222.3 | 165.1 | 2.77 | 20 | 16 |
| <sup>13</sup>C6-Carbaryl (carbamate surrogate) | 208.2 | 151.1 | 2.81 | 25 | 13 |
| Carbaryl | 202.2 | 145.1 | 2.81 | 30 | 16 |
| Methiocarb | 226.3 | 169.1 | 3.16 | 30 | 16 |
| ***Organophosphates*** |  |  |  |  |  |
| Acephate | 184.2 | 143.1 | 2.07 | 15 | 13 |
| Oxydemeton methyl | 247.1 | 169 | 2.08 | 20 | 28 |
| Dimethoate | 230.2 | 199.1 | 2.36 | 20 | 13 |
| Naled | 398.1 | 127 | 2.98 | 15 | 25 |
| Azinphos methyl | 318.1 | 132.1 | 3.13 | 15 | 22 |
| Phosmet | 318.3 | 160.1 | 3.17 | 20 | 22 |
| Methyl Parathion | 263.9 | 232.1 | 3.25 | 15 | 22 |
| Malathion | 331.1 | 127 | 3.41 | 15 | 16 |
| Bensulide | 398.2 | 158 | 3.82 | 15 | 34 |
| Diazinon | 305.2 | 169 | 4.1 | 50 | 31 |
| Chlorpyrifos methyl | 322.2 | 125.1 | 4.24 | 45 | 25 |
| D10-Chlorpyrifos (organophosphate surrogate) | 360.2 | 198.1 | 5.05 | 15 | 34 |
| Chlorpyrifos | 350.2 | 198.1 | 5.07 | 20 | 25 |
| ***Pyrethroids*** |  |  |  |  |  |
| Cyfluthrin | 451.2 | 206 | 5.6 | 15 | 34 |
| Cypermethrin | 433.3 | 191 | 5.7 | 15 | 19 |
| L-Cyhalothrin | 467.2 | 225 | 5.7 | 25 | 25 |
| Deltamethrin | 523 | 506 | 5.83 | 30 | 13 |
| Tau Fluvalinate | 503.5 | 208.2 | 6.14 | 50 | 16 |
| Permethrin (trans) | 408.2 | 183 | 6.15 | 15 | 25 |
| <sup>13</sup>C6-trans Permethrin (pyrethroid surrogate) | 414.2 | 189.1 | 6.18 | 15 | 25 |
| Bifenthrin | 440.2 | 181.2 | 6.77 | 15 | 22 |
| ***Triazines*** |  |  |  |  |  |
| Atrazine | 216.2 | 174.2 | 2.91 | 15 | 25 |
| ***Neonicotinoids*** |  |  |  |  |  |
| Imidacloprid | 256.2 | 209.1 | 2.31 | 15 | 25 |
| ***Instrument internal standards*** |  |  |  |  |  |
| CUDA | 341.3 | 216.2 | 3.31 | 15 | 22 |
| PUHA | 251.2 | 114.1 | 2.4 | 15 | 22 |

### Reference material for method development

The reference material used for method development was from pooled MARBLES participants. The pooled sample was thawed on wet ice, vortexed, and 0.5mL volumes were aliquoted into 2mL polypropylene tubes. Samples were stored at -80°C until analysis.

Samples collected from 5 mothers enrolled in the UCD Lactation Study on day 249 were pooled towards the end of the study (when we ran out of MARBLES reference milk), and used to measure analytical reproducibility of the optimal method, as described below.

All experiments were conducted under amber light conditions to avoid potential photo-degradation of compounds. Samples were kept chilled on ice throughout the entire extraction.

### Extraction Methods

As outlined in the Introduction, three extraction methods were attempted. Method 1 tested microwave-assisted digestion of breast milk in acid or base, followed by SPE purification of pesticide analytes with Oasis HLB columns (Waters, Milford, MA, USA). Method 2 was based on a published and validated method for pyrethroids, which utilized a double liquid-liquid extraction with 20 mL hexane:dichloromethane (2:1) followed by dual column purification with alumina and C18 SPE columns.^18^ Method 3 tested double liquid-liquid extraction with low (2 mL) and high (20 mL) volume hexane:dichloromethane (2:1, v/v/), without the subsequent SPE steps, because we realized that analyte recoveries for many compounds were low after using SPE in Methods 1 and 2 (see Results).

#### Method 1: Microwave digestion in acid or base followed by SPE

Breast milk is enriched with lipids in the form of esterified fatty acids,^35^ which can co-extract with pesticides and cause ion suppression during mass-spectrometry analysis.^36^ We therefore tested whether hydrolyzing these lipids would improve pesticide recovery from small volumes (100 µL) of spiked breast milk, by reducing ion suppression. Microwave-assisted hydrolysis in methanolic acid or base was used, in view of recent data by our group showing the rapid break-down of lipid ester bonds in plasma with microwave-assisted digestion.^37^ Methanolic acid and base were used to determine which reagent efficiently breaks lipid ester bonds during microwave-assisted digestion. It was hypothesized that the degradation of complex lipids in milk with this process would generate free fatty acid methyl esters or free fatty acids that elute separately from pesticides on the LC column, thus improving the analyte signal.

One-hundred µL of the reference breast milk or LCMS-grade water (as negative control) were aliquoted into Teflon MarsXpress (PFS) 20mL tubes (CEM, Matthews, NC) containing 50µL of a 1000nM standard pesticide mixture in methanol. Two-hundred microliters of 10% HCl in methanol (v:v) or 200µL of a 3% sodium carbonate base solution in methanol-water (1:1 v/v) were added to the test-tubes. An additional 50µL methanol was added for a final volume of 400µL in each sample. Thus, the HCl and sodium carbonate concentrations amounted to a final concentration of 5% and 1.5%, respectively.

Microwave-assisted digestion was conducted at 122°C for 5 minutes, held for 3 minutes, and finished with a 7-minute cool-down period at variable power, to hold the desired temperatures. Acid and base digests were neutralized with 20uL 1M sodium hydroxide or 25µL (17.4M) glacial acetic acid, respectively. The samples were decanted into 60mg Oasis HLB SPE columns (Waters, Milford, MA, USA) that had been pre-cleaned with one column volume of ethyl acetate and one column volume of methanol, and preconditioned with two column volumes of SPE buffer (5% methanol in LCMS grade water). The tubes were rinsed with an additional 1.5mL of SPE buffer and decanted into the SPE columns. The milk digests were extracted by gravity elution. Light vacuum (∼10mm Hg) was applied when necessary to assist the elution. The columns were then washed with one column volume (∼ 3 mL) of SPE buffer and dried under -15 psi vacuum for 10 minutes. Analytes were eluted with 0.4 mL methanol followed by 1.5 mL ethyl acetate into a 2mL amber glass autosampler vial containing 10uL of 20% glycerol in methanol. The extracts were brought to dryness by centrifugal vacuum with an EZ-2 Plus Series Genevac (SP Scientific, Warminster, PA) for 30 minutes. The residues were reconstituted in 100µL of 200nM CUDA/PHAU internal standard solution, vortexed for 30 seconds at room temperature and chilled in wet ice for 15 minutes. The extracts were transferred to 0.1um Millipore Duropore PVDF centrifugal filters (cat # UFC30VV00; Cork, IRL), centrifuged for 2 minutes at 4500g and 4°C then transferred to a 150µL glass insert in a 2mL amber auto-sampler vial with a slit cap (Waters Corp, Milford, MA), and analyzed by ultra-high performance liquid chromatography coupled to tandem mass-spectrometry (UPLC-MS/MS) as described below. The analyte spike solution was diluted 10x and measured to calculate analyte recoveries (final concentration of 100nM). Surrogate recoveries were determined against the calibration curve standard concentrations (200nM).

#### Method 2: Liquid-liquid extraction with hexane:dichloromethane (2:1 v/v) followed by SPE

In Method 2, we attempted a published procedure which had been validated for human breast milk pyrethroids, to test whether it could also extract organophosphates and carbamates (alongside pyrethroids).^18^ The method involves liquid-liquid extraction followed by two clean-up steps involving alumina and C18 SPE columns. The alumina column traps polar compounds while eluting relatively non-polar pesticides from the liquid-liquid extraction step when acetonitrile is added to the column. Pesticides are then loaded onto a C18 column which traps them while eluting polar compounds (e.g. sugars). The pesticides are eluted from the C18 column with acetonitrile, residues are dried and reconstituted in methanol prior to UPLC-MS/MS analysis. The experimental design was as follows:

1. Human milk spiked with deuterated surrogate standards (n=1) to quantify pesticide background in the milk matrix
2. Water with deuterated surrogate spike (n=1), to quantify pesticide background in the water matrix
3. Human milk spiked with deuterated surrogates and all analytes (n=3), to determine spike recoveries
4. C18 Hypersep breakthrough (i.e. capture of waste prior to elution of pesticide residues) was collected and extracted by liquid-liquid (n=3) to assess losses due to lack of sorbtion on the C18 Hypersep column.

Ten µL of 2000 nM labelled pesticide surrogate standards and ∼1000 nM unlabeled pesticide analyte standard mix were added to 50mL borosilicate glass tubes. One mL of reference breastmilk (n=3) was added to the tubes and vortexed to mix. To determine whether recoveries were affected by matrix effects, labeled surrogates were spiked to 1 mL of milk or water (n=1 each).

Twenty mL of hexane:dichloromethane (2:1 v/v) were added and the tubes were capped and placed in an ultrasonic bath for 15 minutes at room temperature. The samples were then vortexed for 30 seconds to assist emulsification of the phases. Tubes were centrifuged at 3500 rcf for 5 minutes at 4° C. The top organic phase was collected in a 50 mL tube and the liquid-liquid extraction was repeated with an additional 20 mL of hexane:dichloromethane (2:1 v/v).

The total extracts were combined, dried under nitrogen, reconstituted in 500 uL isopropanol:acetonitrile (1:5), vortexed 30 seconds, and loaded onto a pre-conditioned 5 gram basic alumina column (Silicycle, cat# spe-aut-0055-20x) which holds onto polar constituents (e.g. sugar, salts, etc.), while allowing the pesticide analytes to flow through. Pesticides loaded onto the alumina columns were eluted with 20 mL acetonitrile. The 20 mL eluent was then loaded onto a 2 gram C18 SPE column (Thermo Scientific Hypersep C18, cat#60108-701) to further clean up the extract. In this step, the C18 column is expected to hold on to pesticides while eluting polar compounds (e.g. sugars). Additionally, the 20 mL acetonitrile applied to the C18 column was collected to determine potential losses due to lack of complete adsorption of pesticides to the C18 column. The collected ‘waste’ was dried under nitrogen, extracted with hexane:dichloromethane (2:1 v/v) liquid-liquid extraction, reconstituted in 100µL methanol and analyzed by UPLC-MS/MS.

Pesticides trapped on the C18 column were eluted with 20 mL acetonitrile. All eluates were dried by nitrogen gas. The dried residues were reconstituted in 100µL of 200nM CUDA/PHAU internal standard methanol solution, vortexed for 30 seconds at room temperature and chilled in wet ice for 15 minutes. The extracts were then transferred to 0.1um Millipore Duropore PVDF centrifugal filters and centrifuged for 2 minutes at 4500g and 4°C, and transferred to a 150µL glass insert in a 2mL amber autosampler vial with a slit cap and analyzed by UPLC-MS/MS (see below). The analyte spike solution was diluted 10x and measured to calculate analyte recoveries (final concentration of 100nM). The surrogate recoveries were determined against the calibration standard curve concentrations (200nM).

#### Method 3: Liquid-liquid hexane:dichloromethane (2:1 v/v) extraction without SPE columns

As described in the Results Section, pesticide recovery was low for several compounds with the Corcellas et al. method.^18^ We hypothesized that this was due to analyte loss in the alumina and/or C18 SPE columns. Thus, a modified version of the method was attempted without the SPE columns, and using high (20 mL) and low (2 mL) double hexane:dichloromethane (2:1 v/v) of 1 mL or 100 µL of milk, respectively, to determine whether reducing milk volumes improves pesticide recoveries. A previous study demonstrated that analyte recoveries were improved when matrix effects were minimized for lipid measurements, by reducing sample volumes.^38^

An experimental matrix of 1mL (high volume) and 100µL (low volume) of pooled MARBLES breast milk (n=4 per volume) or LCMS-grade water (n=2 per volume) were extracted twice in 20 mL (Method 3a) or 2 mL (Method 3b) of 2:1 hexane:dichloromethane, as described below. The method was also tested at 200 µL milk with the 2 mL low solvent volume to determine whether the pesticide signal could be improved when the milk volume was doubled from 100 µL (Method 3c). Methods 3a and 3b were carried out and reported as one experiment, but are described separately below to allow for inclusion of technical details in each protocol.

#### Method 3a - High-volume double extraction in 20 mL hexane:dichloromethane

Ten microliters of 2000 nM class-specific stable isotope surrogates and ∼1000nM unlabeled pesticide analytes were spiked into hexane-rinsed 50mL borosilicate screw-threaded conical glass tubes, to which 1 mL of pooled breastmilk (n=4) or water negative control (n=2) were added. Contents were vortexed for approximately 2 seconds. Twenty milliliters of 2:1 (v/v) hexane:dichloromethane were added to all tubes. The tubes were capped with Teflon-lined caps, sonicated for 15 minutes and vortexed for 3 minutes. The samples were centrifuged at 3500 rcf (g) at 4°C for 15 minutes to separate the phases. The top hexane:dichloromethane layer was transferred to a second tube and the extraction repeated. The supernatant of the second extraction was pooled with the first one. Total supernatants were brought to dryness by nitrogen evaporation. The residues were reconstituted in 100µL of 200nM CUDA/PHAU internal standard solution, vortexed for 3 minutes at room temperature and chilled in wet ice for 15 minutes. The extracts were transferred to 0.1um Millipore Duropore PVDF centrifugal filters (cat # UFC30VV00), centrifuged for 2 minutes at 4500g and 4°C, transferred to a 150µL glass insert in a 2mL amber autosampler vial with a slit cap (Waters Corp, Milford, MA), and analyzed by UPLC-MS/MS.

#### Method 3b - Low-volume double extraction in 2 mL hexane:dichloromethane

Ten microliters of 2000nM class specific stabile isotope surrogates and ∼1000 nM analyte spike mixture were spiked into hexane-rinsed 13 x 100 mm glass tubes with polypropylene screw-top caps. One-hundred microliters of pooled breastmilk (n=4) or water (n=2) were added and contents were vortexed for 2 seconds. Two milliliters of a 2:1 hexane:dichloromethane solution were added to all tubes, which were then capped and sonicated for 15 minutes then, vortexed for 3 minutes. Samples were centrifuged at 3500 rcf (g) at 4°C for 15 minutes to separate the phases. The top layer was transferred with a glass Pasteur pipette to a second clean tube and the extraction was repeated, adding supernatant to the second vial. Total supernatants were brought to dryness by centrifugal vacuum. Residues were reconstituted in 100µL of 200nM CUDA/PHAU internal standard solution, capped and vortexed for 3 minutes at room temperature, and chilled in wet ice for 15 minutes. Extracts were transferred to 0.1um Millipore Duropore PVDF centrifugal filters, centrifuged for 2 minutes at 4500g and 4°C, then transferred to a 150µL glass insert in a 2mL amber autosampler vial with a slit cap (Waters Corp, Milford, MA), and analyzed by UPLC-MS/MS.

#### Method 3c - Final optimized low-volume double extraction method in 2 mL hexane:dichloromethane

The low-volume protocol was further optimized to increase pesticide yield from breast milk. Briefly, 200µL instead of 100µL of breastmilk or water blank were extracted to test whether increasing the milk volume would increase the analyte signal. Then, as described above (for Method 3b), ten microliters of 2000nM class specific stabile isotope surrogates were spiked into hexane-rinsed 13 ⨯ 100 mm glass tubes with polypropylene screw-top caps. Two-hundred microliters of homogenized breastmilk were added and contents were vortexed for 2 seconds. Reagent blanks consisted of a water matrix, instead of milk. Two mL of a 2:1 hexane:dichloromethane solution were added to all tubes, which were then capped and vortexed for 6 minutes. Samples were centrifuged at 3500 rcf (g) at 4°C for 15 minutes to separate phases. The top layer was transferred with a glass Pasteur pipette to a second clean tube and the extraction was repeated. The supernatant was combined with the first extract. Total supernatants were brought to dryness by centrifugal vacuum. Residues were reconstituted in 100µL of 200nM CUDA/PHAU internal standard solution, capped and vortexed for 3 minutes at room temperature then, chilled in wet ice for 15 minutes. Extracts were transferred to 0.1um Millipore Duropore PVDF centrifugal filters, centrifuged for 2 minutes at 4500g and 4°C, and transferred to a 150µL glass insert in a 2 mL amber autosampler vial with a slit cap (Waters Corp, Milford, MA), and analyzed by UPLC-MS/MS. For all liquid-liquid trials the analyte spike solution was diluted 10x and measured to calculate analyte recoveries (final concentration of 100nM), and surrogate recoveries were determined against the calibration standard concentrations (200nM).

### Analytical Reproducibility

The intra-experimental variability was determined by pooling samples from 5 subjects collected on day 249 (from the UCD Lactation Study) and measuring pesticides in four 200 µL aliquots extracted twice with 2 mL 2:1 v/v hexane:dichloromethane, as described in Method 3c above.

#### UPLC-MS/MS acquisition method

An 8-minute reverse-phase acquisition method was optimized for detecting 31 pesticides, 4 class specific stable isotopes, and 2 internal standard instrument controls by manual infusion using positive mode electrospray ionization. Analytes were resolved and detected with a Shimadzu Nexera 30AD UPLC coupled to an API Sciex 6500 triple quadrupole mass spectrometer (Sciex, Redwood City CA). Optimized analyte precursor and product ions, declustering potentials, collision energies, and retention times are shown in **Table 1**. Global source parameters were optimized for sensitivity by injection over the UPLC gradient on the API Sciex 6500 in multiple reaction monitoring mode using the instrument settings shown in **Supplemental Table S3**. Pesticides were separated on a Shimadzu 30AD UPLC system in 8 minutes at a flow rate of 0.350mL per minute on a 2.1 x 150mm, 2.7µm Ascentis Express C18 column (Supelco) fitted with a 0.2-micron stainless steel guard column (Waters Corp), at 35°C. The mobile phases were A: 10mM ammonium formate and 0.1% formic acid in LCMS grade, 0.2 micron filtered Optima Water, and B: 10% isopropanol in LCMS grade acetonitrile, 0.2 micron filtered. The UPLC gradient and instrument module parameters are presented in **Supplemental Table S4**.

#### Calculations

Limits of detection (LOD) and limits of quantification (LOQ) were estimated according to the Environmental Protection Agency (EPA) method (40 CFR, Appendix B to Part 136 revision 1.11, U.S. and EPA 821-R-16-006 Revision 2, Procedure 1.c). Specifically, 1-tailed t-tests were run between successive concentrations of calibration standards (n=3 per standard concentration) to determine the region of the calibration where a significant change in sensitivity occurred (p<0.05), ‘i.e., a break in the slope of the calibration’. The standard deviation (σ) of the first significantly different calibration standard replicates was used to estimate the LOD, and back-calculated to sample concentration in nM (sample concentration multiplier). Using the Students t-Distribution, the t-value was determined at both a 95% and 99% 1-tail confidence level to define the LOD and LOQ such that:

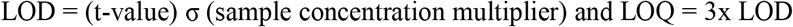

The unlabeled pesticide analyte spike recoveries were calculated as follows:

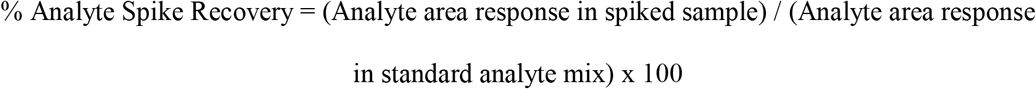

The labeled surrogate spike recoveries were calculated as follows:

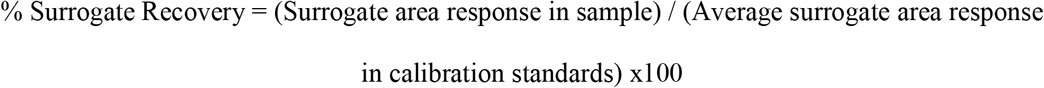

The internal standard recovery, measuring instrument performance across a run of samples and between batches, was calculated at follows:

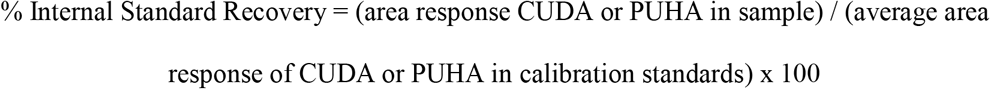

Matrix-corrected surrogate spike recoveries were calculated in study samples to probe for matrix effects between study batches, using area response ratios with the CUDA or PUHA internal standards. Recoveries were calculated as follows:

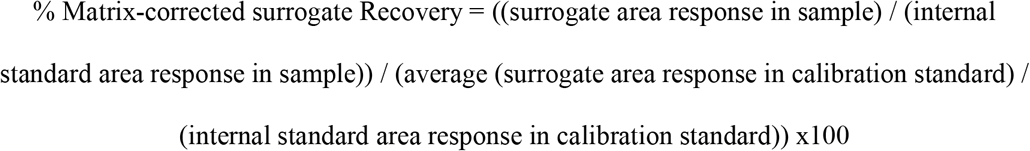

Intra-experimental variability was calculated as the % coefficient of variation (CV) as follows:

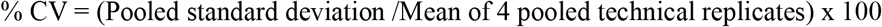

#### Statistical and data analysis

All study sample pesticide residues were quantified and analyzed by calibration curves using area ratios, with their respective labelled class surrogates, on AB Sciex MultiQuant v. 3.1 software (Sciex, Redwood City). Standard curves were fitted with a quadratic regression function and 1/x weighing.^39^ Breast milk samples collected from 79 women were analyzed for pesticide content and compared to LOD and LOQ values at 95% and 99% Confidence Interval (CI).

An unpaired t-test was used to compare analyte recoveries from 1 mL versus 0.1 mL breast milk. Statistical significance was set at P < 0.05.

## Results

### UPLC-MS/MS Method performance

We incorporated 31 pesticides listed in **Table 1** for detection by UPLC-MS/MS. The parent ion mass, product ion mass, retention time, declustering potential and collision energy for each pesticide is presented in the table. Reliable signals for esfenvalerate and methidathion were not obtained with the UPLC-MS/MS conditions listed in **Table 1**, and therefore excluded from the assay. As will be presented below, acephate had a low extraction efficiency of <6% with all methods tested, so it was dropped from the assay when the UCD Lactation Study samples were measured. Thus, the final method incorporated 28 analytes.

### Microwave digestion (Method 1)

Microwave-assisted extraction in 5% HCl or 1.5% sodium carbonate resulted in low recoveries of pesticides spiked to 100 µL of reference breast milk and water. As shown in **Table 2**, the percent recovery for most compounds after acid or base digestion was below 60%, and in many cases, ranged between 0 to 6% in both water and milk. Exceptions were bensulide and deuterated chlorpyrofos (D10-chlorpyrifos), which had milk recoveries of 86% and 93% following base and acid treatment, respectively. For most compounds, the percent recovery from water was comparable to the recovery from breast milk, suggesting that the low pesticide percent recoveries in both matrices were likely due to degradation during microwave-assisted extraction, rather than ion suppression (i.e. matrix effects).

**Table 2.** Mean percent recovery of pesticides from 100 µL breastmilk subjected to microwave-assisted acid or base hydrolysis and extracted with C18 solid phase (n=1 per condition per matrix).

|  | Milk |  | Water |  |
| --- | --- | --- | --- | --- |
|  | 5% HCl | 1.5% Na<sub>2</sub>CO<sub>3</sub> | 5% HCl | 1.5% Na<sub>2</sub>CO<sub>3</sub> |
| *Carbamates* |  |  |  |  |
| <sup>13</sup>C6-Carbaryl | 1% | 0% | 1% | 0% |
| Bendiocarb | 0% | 0% | 0% | 0% |
| Carbaryl | 45% | 23% | 10% | 9% |
| Carbofuran | 0% | 0% | 0% | 0% |
| Formetanate HCl | 2% | 4% | 1% | 1% |
| Methiocarb | 0% | 0% | 0% | 0% |
| Methomyl | 1% | 1% | 0% | 0% |
| <sup>13</sup>C2 <sup>15</sup>N-Methomyl | 0% | 0% | 0% | 0% |
| Oxamyl | 0% | 0% | 0% | 0% |
| Propoxur | 1% | 0% | 0% | 0% |
| *Organophosphates* |  |  |  |  |
| Acephate | 0% | 0% | 1% | 3% |
| Azinphos methyl | 0% | 1% | 0% | 0% |
| Bensulide | 77% | 86% | 26% | 78% |
| Chlorpyrifos | 21% | 18% | 6% | 28% |
| **D10-Chlorpyrifos** | 93% | 61% | 29% | 33% |
| Chlorpyrifos methyl | 12% | 25% | 0% | 4% |
| Diazinon | 1% | 38% | 4% | 33% |
| Dimethoate | 0% | 0% | 0% | 0% |
| Malathion | 0% | 1% | 0% | 0% |
| Methyl Parathion | 59% | 10% | 16% | 65% |
| Naled | 20% | 44% | 17% | 17% |
| Oxydemeton methyl | 0% | 0% | 0% | 0% |
| Phosmet | 0% | 0% | 0% | 0% |
| *Pyrethroids* |  |  |  |  |
| <sup>13</sup>**C6-trans Permethrin** | 9% | 20% | 1% | 1% |
| Bifenthrin | 6% | 4% | 0% | 13% |
| Cyfluthrin | 5% | 3% | 3% | 3% |
| Cypermethrin | 3% | 19% | 3% | 2% |
| Deltamethrin | 1% | 7% | 1% | 1% |
| L-Cyhalothrin | 59% | 21% | 68% | 41% |
| Permethrin (trans) | 11% | 7% | 2% | 2% |
| Tau Fluvalinate | 0% | 5% | 0% | 0% |
| *Triazine* |  |  |  |  |
| Atrazine | 0% | 47% | 0% | 52% |
| *Neonicotinoid* |  |  |  |  |
| Imidacloprid | 10% | 12% | 75% | 13% |

### Liquid-liquid extraction followed by SPE clean-up (Method 2)

**Table 3** shows the percent recoveries of pesticide analytes and/or surrogates spiked to 1mL water or breast milk, extracted twice with 20 mL of 2:1 dichloromethane/hexane, and subjected to alumina Silicycle and C18 Hypersep clean-up.^18^ The first two columns of the table show the recoveries from water and milk spiked with the four deuterated surrogates only. In general, surrogate standard recoveries were low. As shown, the percent recovery of ^13^C2^15^N-Methomyl and ^13^C6-Carbaryl was 12-17% in water and milk matrices. The recovery of D10-Chlorpyrifos was 14% in water and 3% in milk; the recovery of ^13^C6-trans Permethrin was 17% in water and 2% in milk. The far lower recoveries in milk (versus water) suggest ion suppression caused by the milk matrix.

**Table 3.** Percent recoveries of labeled and unlabeled pesticide spikes from 1mL water or breast milk extracted twice with 20 mL of 2:1 dichloromethane/hexane, followed by alumina Silicycle and C18 Hypersep column clean-up.

| Pesticide Analytes | Spike recoveries over entire method |  |  | Hypesep C18 losses in the milk matrix |
| --- | --- | --- | --- | --- |
|  | Water with Surrogate Spike only (n=1) | Milk with Surrogate Spike only (n=1) | Ave Milk with Analyte Spikes (n=3) | Ave losses of analytes in milk through C18 column (n=3) |
| *Carbamate* |  |  |  |  |
| Formetanate HCl | 0% | 1% | 9.13% ± 0.39% | 2.23% ± 0.038% |
| Oxamyl | 0% | 0% | 21.3% ± 0.4% | 56.2% ± 0.053% |
| Methomyl | 0% | 0% | 45.1% ± 0.26% | 64.7% ± 0.14% |
| **<sup>13</sup>C2<sup>15</sup>N-Methomyl** | **17%** | **14%** | **24.8% ± 2.9%** | **63.9% ± 1.4%** |
| Bendiocarb | 0% | 0% | 10.9% ± 0.0082% | 63% ± 0.11% |
| Carbofuran | 0% | 0% | 14% ± 0.045% | 78% ± 0.17% |
| Propoxur | 0% | 0% | 14.4% ± 0.046% | 77.8% ± 0.18% |
| Carbaryl | 0% | 0% | 13.3% ± 0.016% | 66.2% ± 0.033% |
| Methiocarb | 0% | 0% | 9.3% ± 0.22% | 44.9% ± 0.67% |
| **<sup>13</sup>C6-Carbaryl** | **13%** | **12%** | **12.9% ± 1%** | **63.2% ± 3.8%** |
| *Organophosphate* |  |  |  |  |
| Oxydemeton methyl | 0% | 0% | 0.674% ± 0.43% | 1.99% ± 1% |
| Acephate | 0% | 0% | 0.119% ± 0.081% | 0.1% ± 0.03% |
| Chlorpyrifos | 0% | 0% | 6.25% ± 0.93% | 22.9% ± 0.48% |
| Dimethoate | 0% | 0% | 10.3% ± 0.0064% | 37.8% ± 0.1% |
| Naled | 0% | 0% | 2.09% ± 1.3% | 0.588% ± 0.41% |
| Azinphos methyl | 0% | 0% | 12.8% ± 4.4% | 49.3% ± 12% |
| Phosmet | 0% | 0% | 13.6% ± 4% | 73.4% ± 21% |
| Methyl Parathion | 0% | 0% | 10.3% ± 3.8% | 36.9% ± 7.1% |
| Malathion | 0% | 0% | 10.3% ± 2% | 53.2% ± 7.6% |
| Bensulide | 0% | 0% | 9.76% ± 1.1% | 56% ± 1.4% |
| Diazinon | 0% | 0% | 0.007% ± 0.0064% | 0.643% ± 0.33% |
| Chlorpyrifos methyl | 0% | 0% | 9.05% ± 0.76% | 40.1% ± 1.2% |
| **D10-Chlorpyrifos** | **14%** | **3%** | **6.39% ± 5.7%** | **25% ± 19%** |
| *Pyrethroid* |  |  |  |  |
| Cyfluthrin | 2% | 8% | 29.8% ± 9.1% | 39% ± 14% |
| Cypermethrin | 0% | 0% | 4.36% ± 0.18% | 20.7% ± 2.1% |
| L-Cyhalothrin | 7% | 6% | 12.6% ± 7.6% | 21.8% ± 7.2% |
| Deltamethrin | 0% | 0% | 4.69% ± 0.66% | 23.9% ± 3.8% |
| Tau Fluvalinate | 0% | 0% | 5.63% ± 0.093% | 20.3% ± 0.82% |
| Permethrin (trans) | 0% | 0% | 5.15% ± 0.53% | 19.3% ± 1.4% |
| Bifenthrin | 0% | 0% | 4.29% ± 0.68% | 17.2% ± 0.63% |
| **<sup>13</sup>C6-trans Permethrin** | **17%** | **2%** | **4.89% ± 3.3%** | **18.3% ± 14%** |
| *Triazine* |  |  |  |  |
| Atrazine | 0% | 0% | 21% ± 0.35% | 39% ± 0.4% |
| *Neonicotinoid* |  |  |  |  |
| Imidacloprid | 0% | 0% | 3.93% ± 0.084% | 23% ± 0.29% |

The third column of **Table 3** shows the percent spike recovery of both unlabeled and deuterated (labeled) pesticides. Standard recoveries for both labeled and unlabeled pesticides were <30% for all classes, with the exception of methomyl, at 45.1%. Acephate, naled, oxydemton methyl, and diazinon had recoveries near zero.

Further analysis of the waste wash collected after 20mL acetonitrile from the alumina column was decanted onto the C18 column, revealed that the low recovery for most compounds was due to losses in the C18 Hypersep column, as shown in the fourth column of **Table 3**. Most carbamates had over 60% recovery from the waste, organophosphates had 23-73% recovery, and pyrethroids, atrazine and imidacloprid had 17-39% recoveries. Thus, all compounds had higher recoveries in waste than through the extraction method itself. Notably, losses in labeled and unlabeled standards were somewhat proportional within each class of compounds, suggesting that both the labeled and unlabeled standards behaved similarly through the columns.

### Liquid-liquid extraction at low and high milk volumes (Method 3a vs 3b)

In Method 3a and 3b, 1 mL and 100 µL of pooled MARBLES breast milk samples (or water blanks) were spiked with labeled and unlabeled pesticide standards, and extracted twice with 20 mL and 2 mL of 2:1 v/v hexane:dichloromethane, respectively. As shown in **Table 4**, pesticide spike recoveries in water were similar at both 1mL and 100uL volumes (n=2 per volume). However, spike recoveries were significantly lower for 6 carbamates, 9 organophosphates, 3 pyrethroids, atrazine and imidacloprid, in 1 mL compared to 100 µL milk (n=4 per volume), suggesting significant matrix effects on pesticide recoveries at high milk volumes. Acephate recovery was between 3 to 6%, irrespective of matrix or matrix volume, indicating a lack of partitioning into the organic phase during liquid-liquid extraction due to its high polarity (K_ow_ = 0.13 at 25°C or Log K_ow_ = -0.85).

**Table 4.** Analyte percent spike-recoveries from 0.1 or 1 mL of MARBLES breast milk or water following 2:1 hexane:dichloromethane liquid-liquid extraction (no SPE clean-up).

| % Spiked Recoveries | Milk |  | p-value Milk<br>1mL vs.<br>100μL | Water |  |
| --- | --- | --- | --- | --- | --- |
|  | 100μL (n=4) | 1 mL (n=4) |  | 100μL (n=2) | 1 mL (n=2) |
| *Carbamates* |  |  |  |  |  |
| Bendiocarb | 104% ± 14% | 76.6% ± 13% | 0.03005 | 133% (140, 130) | 109% (110, 110) |
| Carbaryl | 114% ± 14% | 94.6% ± 9.1% | 0.05708 | 114% (110, 120) | 115% (120, 110) |
| **<sup>13</sup>C6-Carbaryl** | **112% ± 11%** | **83.2% ± 9.2%** | **0.006882** | **121% (110, 130)** | **96.8% (96, 98)** |
| Carbofuran | 102% ± 2.8% | 69.8% ± 11% | 0.001352 | 109% (110, 110) | 104% (110, 100) |
| Formetanate HCl | 105% ± 3.6% | 68.3% ± 12% | 0.0009999 | 115% (110, 120) | 111% (110, 110) |
| Methiocarb | 83.9% ± 4.9% | 57.8% ± 5.7% | 0.0004274 | 99% (94, 100) | 106% (110, 110) |
| Methomyl | 89.7% ± 5.6% | 56.4% ± 12% | 0.002614 | 94.8% (95, 94) | 89.9% (91, 89) |
| **<sup>13</sup>C2<sup>15</sup>N-Methomyl** | **75.2% ± 6%** | **51.3% ± 6.4%** | **0.001627** | **87.6% (92, 83)** | **76.9% (74, 79)** |
| Oxamyl | 124% ± 11% | 111% ± 9.9% | 0.1115 | 111% (110, 110) | 136% (110, 170) |
| Propoxur | 102% ± 11% | 62.4% ± 14% | 0.00356 | 106% (110, 100) | 85.8% (93, 79) |
| *Organophosphates* |  |  |  |  |  |
| Acephate | 9.67% ± 1.7% | 8.58% ± 0.58% | 0.2759 | 9.22% (8.5, 9.9) | 6.26% (4.8, 7.7) |
| Azinphos methyl | 103% ± 33% | 95.2% ± 11% | 0.6765 | 118% (110, 130) | 92% (84, 100) |
| Bensulide | 103% ± 8.7% | 33.7% ± 3.8% | 0.000006374 | 87.2% (79, 96) | 78.2% (85, 71) |
| Chlorpyrifos | 47.3% ± 5.3% | 10.8% ± 2.2% | 0.00001505 | 91.1% (84, 99) | 90.3% (84, 96) |
| **D10-Chlorpyrifos** | **37.9% ± 6.2%** | **7.41% ± 1.3%** | **0.00007154** | **80.5% (80, 81)** | **77.1% (74, 80)** |
| Chlorpyrifos methyl | 82.8% ± 6.4% | 34.9% ± 4.1% | 0.00001575 | 76% (72, 80) | 77.2% (72, 83) |
| Diazinon | 76.9% ± 7.3% | 28% ± 3.5% | 0.00002046 | 101% (96, 110) | 81.6% (82, 81) |
| Dimethoate | 51.8% ± 3.3% | 44.7% ± 3.5% | 0.0262 | 50.7% (49, 53) | 43.4% (43, 44) |
| Malathion | 87% ± 13% | 19.5% ± 4.7% | 0.00006147 | 109% (100, 120) | 105% (96, 110) |
| Methyl Parathion | 97.9% ± 4.2% | 92.8% ± 8.7% | 0.3292 | 96.2% (94, 98) | 96.6% (83, 110) |
| Naled | 49.4% ± 7.5% | 27.4% ± 3.4% | 0.001787 | 92.8% (87, 99) | 42.2% (47, 37) |
| Oxydemeton methyl | 49.5% ± 4.8% | 26.2% ± 3% | 0.0001633 | 49.6% (46, 53) | 36.6% (26, 47) |
| Phosmet | 253% ± 30% | 413% ± 66% | 0.0046 | 142% (140, 150) | 127% (140, 110) |
| *Pyrethroid* |  |  |  |  |  |
| Cyfluthrin | 128% ± 37% | 121% ± 39% | 0.8048 | 75.3% (81, 70) | 82.7% (77, 89) |
| Cypermethrin | 61.2% ± 31% | 32.1% ± 2.9% | 0.1092 | 142% (140, 150) | 121% (110, 130) |
| Deltamethrin | 65.6% ± 28% | 16.4% ± 4.3% | 0.01277 | 174% (160, 190) | 144% (140, 150) |
| L-Cyhalothrin | 67.8% ± 36% | 45.4% ± 12% | 0.2854 | 138% (170, 110) | 119% (130, 110) |
| Permethrin (trans) | 64.2% ± 17% | 30.1% ± 5.1% | 0.009371 | 238% (240, 240) | 169% (160, 180) |
| <sup>13</sup>C6-trans Permethrin | 45.3% ± 13% | 6.18% ± 0.89% | 0.0009599 | 209% (210, 200) | 162% (160, 170) |
| Tau Fluvalinate | 66.6% ± 15% | 11% ± 3.5% | 0.0003182 | 207% (210, 210) | 183% (170, 200) |
| *Triazine* |  |  |  |  |  |
| Atrazine | 90.4% ± 9.6% | 57.3% ± 3.9% | 0.0006892 | 106% (100, 110) | 94.9% (91, 99) |
| *Neonicatinoid* |  |  |  |  |  |
| Imidacloprid | 89.3% ± 5.5% | 74.2% ± 6% | 0.009891 | 90.7% (88, 93) | 72.8% (59, 87) |

Pooled MARBLES milk samples (100 uL and 1 mL) were spiked with labeled surrogate standards only to quantify pesticide levels in this cohort following liquid-liquid extraction (n=4 replicates per volume). We expected to observe higher pesticide concentrations with 100 uL compared to 1 mL milk, based on our observations of higher analyte and surrogate spike recoveries in 100 uL of milk (**Table 4**). Indeed, as shown in **Table 5**, pesticide concentrations were significantly higher for most analytes detected above the 95% LOD, at 100 µL compared to 1 mL milk (99% LODs are also provided for reference in the table). The only exception was azinphos methyl, which was 1.98 nM in 100 µL and 5.16 in 1 mL breast milk (P<0.05). Two carbamates, carbofuran and methomyl, were observed at 100 µL, but were not detected at 1 mL. Deltamethrin (pyrethroid) and atrazine (triazine) were also seen at 100 µL but not at 1 mL. Overall, these data confirm our findings from the spike recovery study (**Table 4**), indicating that less milk volume increases pesticide detectability and measured concentrations in milk.

**Table 5.** Average of pesticide concentrations (nM) quantified and above LOD at 95% CI in 100 µL and 1 mL of MARBLES pooled breast milk (n=4 replicates), and assosiated water blank values. ND indicates not detected or below the LOD at 95% CI.

| Mean Analytes above LOD at 95% CI (nM) | 100 μL MARBLES Breast milk (n=4) | CV | 1 mL MARBLES Breast milk (n=4) | CV | t-test (95% CI) | 100 μL Water blank mean (range) (n=2) | 1 mL Water blank (n=1) |
| --- | --- | --- | --- | --- | --- | --- | --- |
| ***Carbamates*** |  |  |  |  |  |  |  |
| Carbaryl | 1.23 ± 0.24 | 19% | 0.427 ± 0.051 | 12% | **0.00008** | 0.296 (0.477, 0.114) | 0.11 |
| Carbofuran | 0.0306 ± 0.0025 | 8% | ND | | | 0.0214 (0.025, 0.0177) | 0.004 |
| Methiocarb | 0.0489 ± 0.036 | 73% | 0.032 ± 0.0044 | 14% | 0.33 | 0.0877 (0.0333, 0.142) | 0.00717 |
| Methomyl | 0.128 ± 0.072 | 57% | ND | | | 0.074 (0.0936, 0.0544) | 0.00586 |
| Oxamyl | 0.707 ± 0.52 | 73% | 0.611 ± 0.035 | 6% | 0.69 | 1.40 (0.479, 2.33) | 0.0185 |
| Propoxur | 0.796 ± 0.15 | 19% | 0.187 ± 0.06 | 32% | **0.00003** | 0.633 (0.452, 0.813) | 0.0637 |
| ***Organophosphates*** |  |  |  |  |  |  |  |
| Azinphos methyl | 1.98 ± 1.3 | 64% | 5.16 ± 1.1 | 22% | **0.0029** | 0.567 (0.278, 0.855) | 0.145 |
| Bensulide | 0.138 ± 0.059 | 43% | 0.0359 ± 0.013 | 35% | **0.0054** | 0.0563 (0.0604, 0.0522) | 0.0141 |
| Diazinon | 0.0465 ± 0.0055 | 12% | 0.00651 ± 0.00088 | 13% | **0.0000002** | 0.0533 (0.0526, 0.0539) | 0.00673 |
| Dimethoate | 0.0251 ± 0.0078 | 31% | 0.00164 ± 0.00017 | 11% | **0.00015** | 0.0129 (0.0139, 0.0119) | 0.0013 |
| Malathion | 0.163 ± 0.07 | 43% | 0.0584 ± 0.017 | 28% | **0.012** | (ND, 0.351) | 0.00269 |
| Oxydemeton methyl | 0.09 ± 0.033 | 37% | 0.0423 ± 0.0024 | 6% | **0.013** | 0.120 (0.0859, 0.154) | 0.0431 |
| Phosmet | 99.3 ± 10 | 10% | 22.1 ± 1.1 | 5% | **0.0000001** | (ND, 90) | 7.53 |
| ***Pyrethroids*** |  |  |  |  |  |  |  |
| Deltamethrin | 2.09 ± 1.4 | 69% | ND | | | 1.49 (0.919, 2.06) | 0.961 |
| Permethrin (trans) | 10.8 ± 2.9 | 27% | 1.74 ± 0.37 | 21% | **0.00013** | 0.792 (0.123, 1.46) | 0.109 |
| ***Triazines*** |  |  |  |  |  |  |  |
| Atrazine | 0.0524 ± 0.017 | 33% | ND | | | 0.0781 (0.0551, 0.101) | 0.00591 |
| ***Neonicotinoids*** |  |  |  |  |  |  |  |
| Imidacloprid | 0.445 ± 0.039 | 9% | 0.345 ± 0.013 | 4% | **0.00056** | 0.0224 (0.0263, 0.0185) | 0.00485 |

We also observed peaks, above the LOD (reported in **Table 6**), in the one or two water blanks extracted with the same protocol as the milk samples (**Table 5**). The blank concentrations were variable and exceeded the concentrations of pesticides measured in milk for methiocarb, oxamyl, diazinon, melathoin, oxydemeton methyl and atrazine at 100 µL and/or 1 mL (**Table 5**). Ideally, a minimum of 3 blanks per assay (instead of 1 or 2) would have provided a more accurate representation of the background to allow for blank subtraction from analyte values. We took this into account when analyzing the UCD Lactation Study samples (below), by better quantifying the background signal and subtracting it from measured pesticide levels.

**Table 6.** Estimated instrument limits of detection (LOD) at confidence intervals (CI) of 95% and 99%, and regression coefficient (R^2^) of the standard curve fit for each of 28 pesticide analytes measured in MARBLES pooled milk samples and the UCD Lactation Study.

| | 95% CI | | 99% CI | | Curve $R^2$ (Quadratic, weighted 1/x) | |
| --- | --- | --- | --- | --- | --- | --- |
| (nM) | MARBLES | Lactation Study | MARBLES | Lactation Study | MARBLES | Lactation Study |
| *Carbamates* |  |  |  |  |  |  |
| Bendiocarb | 0.051 | 0.031 | 0.121 | 0.054 | 0.997 | 0.984 |
| Carbaryl | 0.040 | 0.117 | 0.096 | 0.205 | 0.999 | 0.976 |
| Carbofuran | 0.008 | 0.057 | 0.019 | 0.101 | 0.999 | 0.982 |
| Formetanate HCl | 3.340 | 0.039 | 7.96 | 0.068 | 0.999 | 0.996 |
| Methiocarb | 0.023 | 0.098 | 0.054 | 0.172 | 1 | 0.981 |
| Methomyl | 0.022 | 0.076 | 0.052 | 0.134 | 0.1 | 0.988 |
| Oxamyl | 0.063 | 0.539 | 0.151 | 0.947 | 0.999 | 0.986 |
| Propoxur | 0.057 | 0.555 | 0.137 | 0.975 | 0.954 | 0.972 |
| *Organophosphates* |  |  |  |  |  |  |
| Azinphos methyl | 0.087 | 1.260 | 0.209 | 2.210 | 0.999 | 0.982 |
| Bensulide | 0.007 | 0.032 | 0.016 | 0.056 | 1 | 0.997 |
| Chlorpyrifos | 0.370 | 0.024 | 0.882 | 0.042 | 1 | 0.998 |
| Chlorpyrifos methyl | 1.88 | 0.022 | 4.48 | 0.039 | 0.992 | 0.993 |
| Diazinon | 0.002 | 0.021 | 0.005 | 0.037 | 1 | 0.999 |
| Dimethoate | 0.001 | 0.046 | 0.002 | 0.081 | 0.999 | 0.994 |
| Malathion | 0.005 | 0.025 | 0.012 | 0.044 | 1 | 0.989 |
| Methyl Parathion | 4.93 | 0.106 | 11.7 | 0.186 | 1 | 0.999 |
| Naled | 3.22 | 0.033 | 7.67 | 0.058 | 0.997 | 0.995 |
| Oxydemeton methyl | 0.013 | 0.073 | 0.032 | 0.128 | 0.999 | 0.990 |
| Phosmet | 1.77 | 1.090 | 4.22 | 1.910 | 0.997 | 0.988 |
| *Pyrethroids* |  |  |  |  |  |  |
| Bifenthrin | 0.716 | 142.0 | 2.73 | 250.0 | 0.986 | 0.949 |
| Cyfluthrin | 39.3 | 716.0 | 103 | 1260.0 | 0.999 | 0.900 |
| Cypermethrin | 8.43 | 93.5 | 20.1 | 164.0 | 0.995 | 0.968 |
| Deltamethrin | 1.03 | 55.1 | 2.45 | 96.8 | 0.998 | 0.990 |
| L-Cyhalothrin | 56.5 | 144.0 | 135 | 253.0 | 0.968 | 0.942 |
| Permethrin (trans) | 1.49 | 47.0 | 3.55 | 82.6 | 0.991 | 0.993 |
| Tau Fluvalinate | 1.53 | 119.0 | 3.64 | 210.0 | 0.993 | 0.969 |
| *Triazine* |  |  |  |  |  |  |
| Atrazine | 0.012 | 0.042 | 0.028 | 0.074 | 0.999 | 0.980 |
| *Neonicotinoid* |  |  |  |  |  |  |
| Imidacloprid | 0.068 | 0.026 | 0.163 | 0.046 | 0.999 | 0.972 |

### Comparison of surrogate recoveries between 100 and 200µL of milk matrix (Method 3c)

Inspection of the matrix-corrected surrogate standard recoveries in the pooled MARBLES samples revealed a relatively low recovery of 34% for ^13^C6-trans Permethrin in 100 µL milk, as shown in the first row of **Supplemental Table 5**. Doubling the volume of milk obtained from pooled samples of the Day 249 UCD Lactation Study increased ^13^C6-trans Permethrin recovery to 84% without markedly affecting the recovery of other surrogate standards (**Supplemental Table 5**, second row). The increase in analyte signal was maintained for the 79 samples from the UCD Lactation Study analyzed by UPLC-MS/MS (**Supplemental Table 5**, third row). Overall, the data suggest that 200 µL milk volume provides an enhanced signal on the mass-spectrometer compared to 100 µL, without causing significant ion suppression as observed in the 1 mL milk volume.

Comparison of labelled surrogate recoveries after correction with the CUDA and / or PUHA internal standards, showed a reduction in the response between the MARBLES and UCD Lactation Study runs, which were separated by a period of 6 months (**Supplemental Table 5**). This is likely due to loss in sensitivity between UPLC-MS/MS runs. It is unlikely due to matrix effects, because as discussed above, surrogate standard recoveries were similar or higher when the milk volume increased from 100 µL to 200 µL.

### Estimated LOD and LOQ

The LOD and LOQ at 95% and 99% CI, were determined by analyzing successive concentrations of calibration standards across all 28 compounds, at the time the method was being developed and tested on MARBLES breast milk, and when the UCD Lactation study samples were analyzed, 6 months after MARBLES. LODs for both cohorts are shown in **Table 6**. LOQs, reported as 3 times the LOD measured values, are in **Supplemental Table 7**.

For the MARBLES study, the LODs at 95% and 99% CI ranged from 0.001 to 56.5 nM and 0.002 to 135nM, respectively across the 28 compounds (**Table 6**). LODs for the UCD Lactation Study were variable: in some cases comparable, higher or lower than MARBLES depending on the analyte, and ranged between 0.021-716 nM and 0.037-1260 nM at 95% and 99% CI, respectively. Higher LODs observed among the late-eluting pyrethroids, are likely due to changes in instrument performance between runs, based on batch-to-batch differences in the CUDA/PUHA internal standard response as shown in **Supplemental Table 5**.

The regression coefficient for each standard curve is also presented in **Table 6**. As shown, the R^2^ value was greater than 0.99, confirming an acceptable goodness of fit for each analyte.

### Intra-experimental variability of pesticide concentrations

Two hundred µL of pooled milk samples (n=4) obtained on day 249 of lactation from the UCD Lactation Study were analyzed with Method 3c, to determine the intra-experimental variability based on the calculated CV. **Table 7** shows mean concentrations of the detected analytes following blank subtraction, and the average blank values in water, relative to the 95% and 99% CI LOD and LOQ. As shown, a total of 21 pesticides were detected at 95% CI, including 8 carbamates, 11 organophosphates, atrazine and imidacloprid. No pyrethroids were detected, likely due to the high LOD at 95% CI.

**Table 7.**
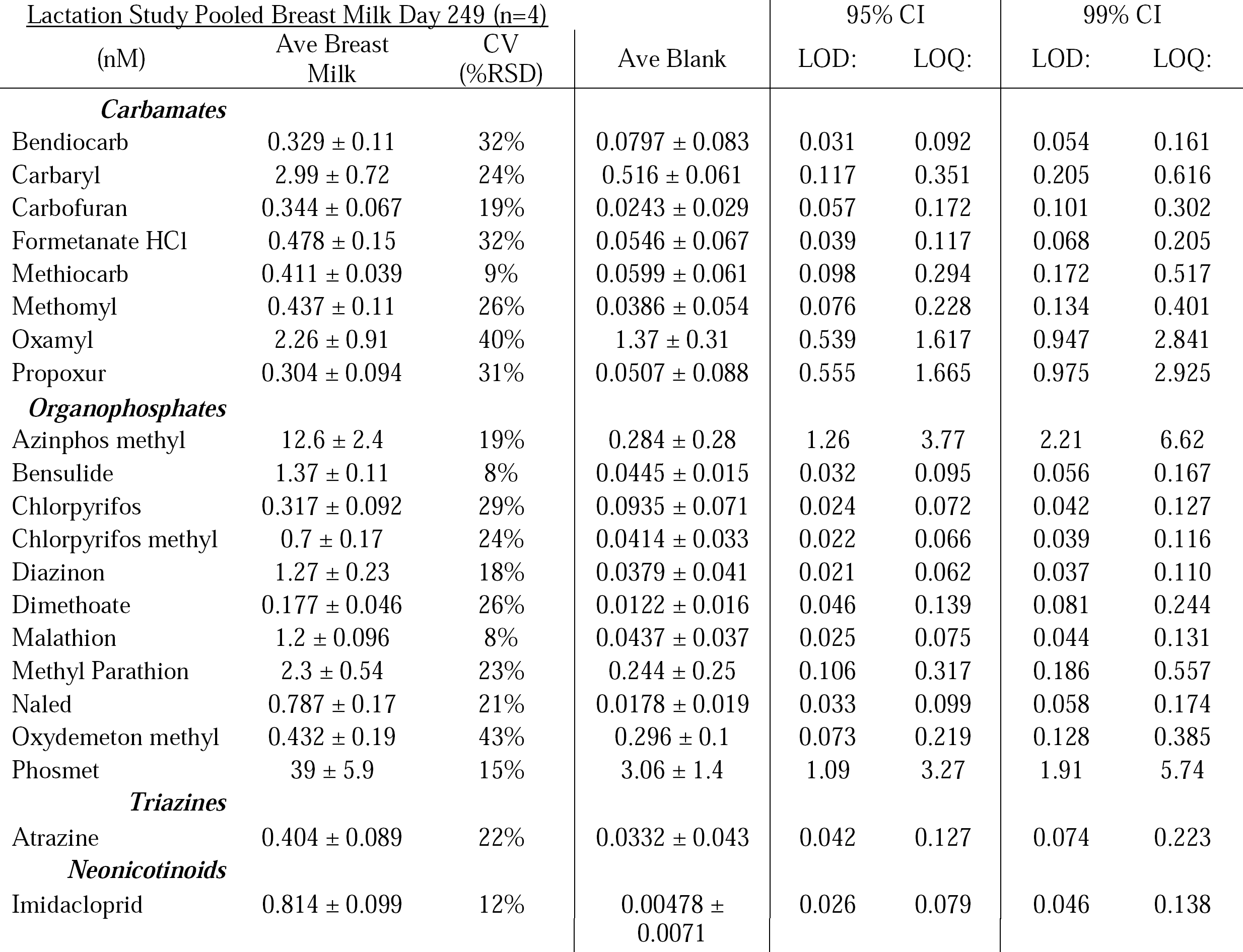
UCD Lactation Study day 249, 200µL pooled breast milk (n=4) blank-subtracted mean concentrations (nM) quantified above LOD (95% CI) with estimated LOD and LOQ at 95% and 99% CI shown on the right.

For detected compounds, the CV was generally below 30%, consistent with the literature.^16, 20^ It ranged between 9-43%. Two compounds had a CV at or above 40%; oxamyl at 40% and oxydemeton methyl at 43%.

### Pesticide concentrations in the UCD Lactation Study

For the UCD Lactation Study (n=79), 200 µL of breast milk were extracted with Method 3c and quantified alongside 3 water blanks, which were subtracted from pesticides measured in the milk samples to account for background noise. Population mean, range and blank values of pesticides, above the 95% CI LOD is reported in **Table 8**. The percentage of pesticides at or above the estimated LOD and LOQ at both 95% and 99% CIs is also reported. **Supplemental Table 6** shows the raw concentration values for each subject. Raw UPLC-MS/MS chromatograms and corresponding standard curves for each pesticide are shown in **Supplemental Figure 1**.

**Table 8.** Blank-subtracted pesticide means, range, average water blank values, and percent of samples above LODs and LOQs for pesticides measured and observed in 200 uL Lactation Study Breast milk samples from 79 subjects on day 42 postpartum.

| <u>Lactation Study Day 42 Population Mean for 79 Subjects (blank subtracted)</u> |  |  |  | <u>% Lactation Study Samples Above LOD/LOQ</u> |  |  |  |
| --- | --- | --- | --- | --- | --- | --- | --- |
| (nM) | Population Mean | Range | Average Blank Value (n=3) | <u>95% CI</u> |  | <u>99% CI</u> |  |
|  |  |  |  | LOD | LOQ | LOD | LOQ |
| *Carbamates* |  |  |  |  |  |  |  |
| Oxamyl | 1.26 ± 0.83 | (0.137,5.10) | 1.37 ± 0.313 | 79% | 23% | 63% | 2% |
| Carbaryl | 1.87 ± 1.6 | (0.011,9.37) | 0.516 ± 0.061 | 96% | 88% | 92% | 80% |
| *Organophosphates* |  |  |  |  |  |  |  |
| Azinphos methyl | 5.85 ± 6.3 | (0.226,34.2) | (0.285,0.567) | 90% | 49% | 70% | 29% |
| Malathion | 0.139 ± 0.15 | (0.003,0.846) | 0.044 ± 0.037 | 85% | 57% | 73% | 38% |
| Oxydemeton methyl | 0.134 ± 0.16 | (0.0001,0.821) | 0.296 ± 0.101 | 53% | 17% | 40% | 7% |
| Chlorpyrifos | 0.0977 ± 0.11 | (0.00003,0.380) | 0.094 ± 0.071 | 66% | 44% | 47% | 31% |
| Diazinon | 0.0623 ± 0.062 | (0.0003,0.314) | (0.032,0.082) | 87% | 29% | 61% | 10% |
| Chlorpyrifos methyl | 0.0518 ± 0.094 | (0.0001,0.576) | 0.041 ± 0.033 | 66% | 16% | 34% | 8% |
| *Pyrethroids* |  |  |  |  |  |  |  |
| Cypermethrin | 25 ± 25 | (1.59,180) | (2.93,3.55) | 1% | 0% | 1% | 0% |
| Permethrin (trans) | 20 ± 25 | (0.616,150.3) | (0.588,1.47) | 7% | 1% | 4% | 0% |
| *Neonicotinoid* |  |  |  |  |  |  |  |
| Imidacloprid | 0.0769 ± 0.13 | (0.001,0.728) | (0.001,0.013) | 61% | 32% | 42% | 6% |

As shown in **Table 8**, a total of 11 pesticides, including 2 carbamates (oxamyl and carbaryl), 6 organophosphates (azinophos methyl, malathion, oxydemeton methyl, chlorpyrifos, diazinon, chlorpyrifos methyl), 2 pyrethroids (cypermethrin and trans permethrin) and the neonicotinoid, imadacloprid, were detected above the 95% and 99% CI LOD. Detection frequencies at the 95% CI for carbamates, organophsphates, pyrethroids and imidacloprid were 79-96%, 53-90%, 1-7% and 61% of the total cohort (n=79), respectively.

LOQ detection frequencies were generally lower, as expected. Pyrethroids were barely detected at LOQ of 95% and 99% CI (0-1%). Other compounds were seen at a frequency of 16-88% at 95% CI, and 2-80% at 99% CI.

Concentrations of most compounds were below 1 nM, except for oxamyl (1.3 nM), carbaryl (1.9 nM) and azinphos methyl (5.9 nM). Concentrations of the two detected pyrethroids, cypermethrin and permethrin, spanned a wide range of 1.6-180 nM for cypermethrin and 0.6-150 nM for permethrin.

## Discussion

In the present study, we developed a simple, two-step method for extracting pesticides from 100-200µL of human breast milk. We demonstrated that the percent recovery of pesticides was significantly improved by lowering both the breast milk volume from 1 mL to 100-200 µL, and the dichloromethane/hexane extraction solvent volume from 20 mL to 2 mL, and by eliminating the SPE clean-up steps typically found in current methods. Pesticides were detected in breast milk of pooled MARBLES and UCD Lactation Study samples (day 249 postpartum). Additionally, eleven pesticides were detected in the UCD Lactation cohort of 79 women on day 42 postpartum, at frequencies of 79-96% for carbamates, 53-90% for organophosphates, 1-7% for pyrethroids and 61% for imidacloprid. Atrazine was not detected in the UCD Lactation Study.

Microwave-assisted hydrolysis in methanolic acid or base followed by SPE purification (Method 1) resulted in poor pesticide spike recoveries of <5% for most compounds in both milk and water (**Table 2**). The low recoveries are likely due to losses in the SPE column or degradation of the compounds during microwave-assisted hydrolysis. Elimination of the SPE step or modification of the microwave cycling parameters and acid / base concentration in methanol may improve pesticide recoveries. As is, however, the method is not appropriate for extracting pesticides.

Method 2 was previously validated for pyrethroids and involved liquid-liquid solvent extraction (40 mL total) followed by two SPE steps.^18, 20^ Using this method, we found that spike recoveries were low (∼10%) for all pesticide classes including pyrethroids, due to losses in the SPE column (**Table 3**). Losses in unlabeled pesticides were proportional to the surrogate standards used in their quantification, which means that absolute concentrations would not be impacted after correcting analytes by the surrogate standard. However, the low percent recoveries are likely to reduce sensitivity, because losses in the SPE column imply less analyte being injected into to the mass-spectrometer. This may affect the detectability of pesticides present at low concentrations.

Removing the two SPE steps in Method 3 resulted in a 5-10 fold increase in extraction recoveries, particularly when only 100 µL (versus 1 mL) of milk was extracted with less solvent (2 mL versus 20 mL; **Table 4**). Increasing the breast milk volume from 100 µL to 200 µL also maintained or improved the signal (**Supplemental Table 5**). The improvement in pesticide extraction recoveries in 100 µL or 200 µL compared 1 mL milk is likely due to the elimination of matrix effects associated with ion suppression. This is supported by our observation that analyte recoveries in 100 µL milk were comparable to water control (i.e. no matrix), but significantly higher than 1 mL milk (**Table 4**). Additionally, measured pesticide concentrations in the pooled MARBLES samples were significantly higher or more detectable in 100 µL compared to 1 mL milk (**Table 5**). Milk is a complex matrix, and its lipid constituents are known to suppress or neutralize the charge of molecular ions at the electrospray mass-spectrometry source through increased viscosity of the nebulized droplet surface, thus inhibiting release of charged ions, complexation with macromolecules, and competition for ionic charge, all effectively lowering the signal reaching the detector (Reviewed in ^40^). Our findings are in agreement with studies that reported increased recoveries of other analytes (e.g. oxidized lipids) after reducing the sample matrix amount.^38, 41^

UPLC-MS/MS analysis revealed the unexpected presence of pesticides in blank LCMS-grade water, extracted in the same manner as the milk (**Table 5**). A similar background signal was previously reported by Hao et al. when pesticides were measured in “nanopure” water on the same type of mass-spectrometer used in our study (QTRAP 6500).^42^ The background signal in water is likely due to the highly sensitive QTRAP 6500 detecting molecular ions generated from non-specific interactions between the water, extraction solvents, column and detector. This is supported by data showing that modifying the multiple reaction monitoring conditions decreased the background noise originating from water blanks.^42^ Thus, the detected pesticides in water blanks is not due to contamination per se, but due to water producing artefact signals on the highly sensitive QTRAP 6500. This is why water blank-subtraction is necessary when measuring pesticides with UPLC-MS/MS, particularly for matrices with high water content such as milk.

LOD values were variable between MARBLES and UCD Lactation Study runs, measured 6 months apart (**Table 6**). The variability in LODs is likely due to changes in analyte ionization efficiencies affecting instrument sensitivity between runs. While most analytes had LOD values below or close to 1nM, the LOD for pyrethroids was above 1 nM in both MARBLES (8.4 to 56.5 nM for cyfluthrin, cypermethrin and lambda-cyhalothrin) and UCD Lactation Study cohorts (47-716 nM for all pyrethroids), suggesting low sensitivity to this class of compounds. A possible contributing factor to the lack of sensitivity may be the form of molecular ion, as pyrethroids were better detected with mass-spectrometry as ammonium adducts.^43^ Although acidified ammonium formate was part of the mobile phase in this study, ammonium adduct formation was reported by others to improve when the mobile phase was buffered to pH 6.8.^44^ Instrument and column performance may also change over time and affect sensitivity, suggesting that LOD and corresponding LOQ estimations should be measured at the time of each analysis. Measuring the LOD and LOQ during each run may allow harmonization across batches.

Intra-sample variability assessed in breast milk pooled from 4 different UCD Lactation Study participants at 249 days postpartumshowed acceptable CVs below 30% for most compounds (**Table 7**) and comparable to the literature.^16, 20^ The CVs were also close to 30% in pooled MARBLES samples, although these were not blank-corrected due to the small number of water blanks analyzed at the time (**Table 5**). Overall, the data suggest acceptable reproducibility within cohorts.

In the UCD Lactation cohort, 21 pesticides were detected on day 249 (**Table 7**) compared to 11 detected on day 42 postpartum (**Table 8**). Pesticides that were observed at both time-points were approximately 2-20 times higher in concentration on day 249 than in day 42, and include two carbamates (oxamyl, carbaryl), 6 organophsphates (azinophos methyl, malathion, oxydemeton methyl, diazinon, chlorpyrifos and chlorpyrifos methyl) and imidacloprid. Oxamyl, carbaryl, azinophos methyl, malathion, oxydemeton methyl and diazinon, were also seen in pooled MARBLES samples at concentrations close to the 42-day UCD Lactation Study samples. These observations should be interpreted with caution, however, because unlike the samples measured on day 42 postpartum (in the Lactation UCD study), the measurements performed in MARBLES and on day 249 of the UCD study were done on pooled rather than individual samples. Thus, they do not incorporate the biological variability between mothers.

Analysis of the UCD Lactation Study breast milk showed the presence of 2 carbamates (oxamyl and carbaryl), 6 organophsphates (azinophos methyl, lamathion, oxydemeton methyl, chlorpyrifos, diazinon and chlorpyrifos methyl), 2 pyrethroids (cypermethrin and permethrin) and imidacloprid. Concentrations were highest but variable for pyrethroids (20-25 nM), followed by carbamates (∼1.3-1.9 nM) and organphsphates (0.05-0.139 nM), and are in general agreement with values reported in the literature.^25, 28^ Additionally, not all mothers were exposed to the same pesticides since carbamates, organophsphates, pyrethroids and imidacloprid, were detected at frequencies of 79-96%, 53-90%, 1-7% and 61%, respectively. Differences in pesticide concentrations and detectability reflect variability in exposures from air, dust, water or food,^1-2, 42, 45^ consistent with another study, which reported wide ranges of pesticides in breast milk obtained from women living in both urban and agricultural communities.^16^ Future studies are needed to better identify sources of exposure in these cohorts.

The detection of pesticides in breast milk does not equate to health risks, particularly given the evidence that breast milk is protective against neurodevelopmental disorders.^46^ The present study was specifically designed to develop methods to allow maternal exposure assessments. The simple method developed could be used in future studies to determine whether reducing maternal exposures further enhances the neurodevelopmental benefits of breast-feeding.^46^

In summary, this study validated a simple dichlormethane/hexane extraction method for measuring pesticides in low volumes of breast milk (100-200 µL), and demonstrated the presence of several pesticide classes in breast milk collected from two cohorts, albeit at very low concentrations. Analytical take-aways of the study are three-fold. First, reducing sample amount and solvent volume, and eleminating SPE purification steps reduces matrix effects, thus improving pesticide spike recovery and reproducibility. Second, background analyte levels should be quantified in a representative blank matrix (e.g. water) and subtracted from pesticides values found in the sample (milk). Third, LOD values must be measured at the same time of the run, to account for changes in instrument response over time. These analytical takeaways may be expanded to other biological matrices such as plasma, to shorten cumbersome protocols and enable routine assessments of pesticide exposure.

## Data Availability

The raw data are provided in the uploaded supplement.

## Abbreviations

CI: confidence interval;
CUDA: 1-cyclohexyl ureido dodecanoic acid;
CV: coefficient of variation;
EPA: Environmental Protection Agency;
IRB: Institutional Review Board;
LCMS: liquid chromatography-mass spectrometry;
LOD: Limits of detection;
LOQ;: limits of quantification;
MARBLES: Markers of Autism Risk in Babies - Learning Early Signs;
NIOSH: National Institute for Occupational Safety and Health;
PUHA: 1-phenyl-ureido3-hexanoic acid;
SPE: solid phase extraction;
UCD: University of California - Davis
UPLC-MS/MS: ultra-high performance liquid chromatography coupled to tandem mass-spectrometry

## Author Contributions

T.P., J.T.S., C.K.W., D.H.B., R.J.S., I.H-P. and A.Y.T. designed the analytical experiments and/or clinical studies. T.P. performed the experiments, analyzed the data, and co-wrote the manuscript with A.Y.T. S.E. contributed to the data analysis. All authors have read and approved the paper.

## Acknowledgement

The authors are grateful to all the women who generously donated milk to support this research, Dr. John Newman (USDA) for providing access to the QTRAP 6500 instrumentation in his lab, and Dr. Bruce Hammock (UCD) for suppling the PUHA internal standard.

## Notes

The authors declare no competing financial interest.

## Ethics Statement

All human subject protocols were approved by the University of California, Davis (UCD) Institutional Review Board (# 225645, 216198 and 887479). Written informed consent was obtained from all participants prior to collection of data or specimens.

